# Vaccines against Covid-19, venous thromboembolism, and thrombocytopenia. A population-based retrospective cohort study

**DOI:** 10.1101/2021.07.23.21261036

**Authors:** Joan-Ramon Laporte, Ermengol Coma, Francesc Fina, Luís García-Eroles, Xavier Vidal, Manuel Medina

## Abstract

**Background:** We aimed at estimating the risk of venous thromboembolism (VTE), thrombocytopenia (TCP), and VTE associated with TCP, by age and sex, after the first dose of both adenovirus vector-based and mRNA-based Covid-19 vaccines, and after the second dose of m-RNA vaccines.

**Methods:** In this population-based retrospective cohort study in the national health care databases in Catalonia, we examined three groups: 1 662 719 people 10 years of age and over vaccinated with the first dose of a Covid-19 vaccine, 622 778 with the second dose, and 190 616 diagnosed of Covid-19 in the same period (between1 January 2021 and 18 April 2021). The rates of various clinical presentations of VTE and TCP were compared with those in the reference population (7 013 040 people served by the health care system in 2019). The two primary outcomes were the observed 21 day rate of a composite variable of cerebral venous sinus thrombosis, mesenteric thrombosis, portal vein thrombosis, or any venous thromboembolism (VTE) associated with thrombocytopenia (TCP), and the rate of any VTE associated with TCP (VTE+TCP). Analyses were standardised by age and sex.

**Results:** The 21 day rate per 100 000 of the primary composite variable was 2.15 in the reference population, 5.65 following the first vaccine dose (standardised difference, 2.53 (95% CI [CI], 1.04 to 4.00), and 7.23 following the second dose (standardised difference, 4.07 (95% CI, 1.43 to 6.70). The event rates of VTE+TCP and of all the secondary variables showed the same patterns.

Excess event rates were higher in men than in women, and they were not especially increased in any particular age group. All Covid-19 vaccines were associated with increased rates of the outcome variables.

Excess event rates were much higher in the Covid-19 cohort (35.60 per 100 000 (95% CI, 26.15 to 45.06).

**Conclusions:** We observed increases of rates of venous thromboembolism in usual and unusual anatomical sites and of thrombocytopenia in recipients of both adenovirus vector and mRNA vaccines against Covid-19. Excess rates were higher in men than in women and they were not particularly elevated in any specific age group.

## Introduction

Only six weeks after approval of VaxZevria® at the end of January, a number of cases of thrombosis in unusual anatomical sites in previously healthy individuals, often associated with thrombocytopenia, were reported from several countries. The affected patients were predominantly young women.

On March 31^st^, the EMA executives acknowledged that they were unable to examine whether the risk was particularly high in young women because they ignored the number of vaccinated individuals by age and sex. Apparently based on spontaneous reporting data, they estimated that the risk of cerebral venous thrombosis after the VaxZevria® vaccine was 5 cases per million doses (one in 200 000 doses).^1^ However, published estimates of incidence have ranged from one in 26 000 in Norway to one in 250 000 in Germany.^2–4^ Regrettably, the risk in people over 65 and the risk associated with other Covid-19 vaccines was not been assessed.

More recently, the EMA has announced additional reviews of reports of thromboembolic events in people who had received the adenovirus vector-based Covid-19 Vaccine Janssen, and the mRNA-based vaccines Comirnaty® (Pfizer/BNT) and Spikevax® (previously COVID-19 vaccine Moderna]).^5^

It is unclear whether mRNA-based vaccines against Covid-19 are also linked to thrombosis or thrombocytopenia. The U.S. Vaccine Adverse Event Reporting System (VAERS) database (last accessed 20 June 2021) contains dozens of reports of unusual site venous thrombosis after vaccination with Comirnaty® or with Spikevax®, and cases have also been reported to the EMA and in the literature.^6,7^ In a (preprint) study in the U.S., the incidence of cerebral venous sinus thrombosis (CVST) and of portal vein thrombosis (PVT) in the two weeks following a diagnosis of Covid-19 was around 10-fold the incidence following an mRNA Covid-19 vaccine (44 per 100 000 compared to 4.5 per 100 000).^8^ Surprisingly, however, the incidence rate of CVST and PVT with the mRNA vaccines was 10 times higher than the incidence of CVST following vaccination with VaxZevria® reported by the EMA (5 cases per million vaccinated people) and than the latest reported incidence of CVST following vaccination with the Ad26.COV2.S Janssen vaccine (0.9 per million).

In summary, evidence of a risk of venous thromboembolic disease associated with adenovirus vector based Covid-19 vaccines is accumulating, but many uncertainties remain. Scarce data are available on the risk in people over 65 years of age, on whether it varies with age or sex, whether it is still increased following the second dose, or how it compares with the risk in Covid-19 patients and between the various vaccine types.

The present study had two objectives. First, to assess the incidence rates of various definitions of venous thromboembolism, thrombocytopenia, and venous thromboembolism associated with thrombocytopenia in the vaccinated population and in patients diagnosed of Covid-19, and to compare these rates with corresponding background rates in the same population, by age and sex. Second, to assess these rates in people vaccinated with the first dose of the first three Covid-19 vaccines which have been available in Catalonia, i.e., Comirnaty®, Spikevax®, and VaxZevria®, and in people who received the second dose of Comirnaty® or Spikevax®.

## Methods

### Data sources

Catalonia has a tax-financed universal health-care system. Since 1990 all contacts with hospital and primary care centres are prospectively collected and registered in a population-based register of minimal basic datasets (*Conjunt Mínim Bàsic de Dades*, CMBD). The CMBD contains doctor recorded diagnoses according to ICD-10 (International Classification of Diseases, 10^th^ revision) and clinical activity of all the health centres in Catalonia. It provides exhaustive and valid information on health morbidity, based on data provided by all health centres.^9^ The CMBD register can be linked with the vaccination register through each citizen’s unique registration number.

### Vaccination against covid-19 and study periods

In Catalonia vaccination against Covid-19 started on 27 December 2020 with Comirnaty®. Vaccination with Spikevax® started on 13 January 2021, with VaxZevria® on 8 February, and with the Janssen Ad26.COV2.S vaccine on 22 April. By order of priority, Comirnaty® and Spikevax® were targeted to nursing home residents, front line health professionals, people with severe dependence, those over 80 years of age, and other high risk groups, such as patients who had received a solid organ or an haematopoietic transplantation, those on cancer chemotherapy, those on replacement treatment for end-stage renal disease, or with a primary immunodeficiency syndrome, with HIV infection and a CD4 cell count below 200/mm^3^, with cystic fibrosis, or older than 40 years with Down syndrome. VaxZevria® was initially targeted to health professionals and other social priority groups between 18 and 55 years of age, except the high risk groups described above. Vaccination with VaxZevria® was temporarily halted between 16 and 22 March. From 30 March its use was extended to those 56 to 65 years old, but administration of the second dose was deferred from 12 to 16 weeks. Therefore, we have no data on the risk of thrombosis in recipients of the second dose of VaxZevria®.

### Study outcomes

We focused on venous thromboembolism (VTE) and thrombocytopenia (TCP) occurring in the 21 days following vaccination against Covid-19 or following a diagnosis of Covid-19. We selected two primary outcome variables, based on those ICD-10 diagnoses or groups of diagnoses which correspond to the cases initially reported through the pharmacovigilance systems. First, the rate of a composite variable of cerebral venous sinus thrombosis (CVST), mesenteric thrombosis (MesT), portal vein thrombosis (PVT), or a simultaneous diagnosis of VTE and TCP (“unusual site VTE or VTE associated with TCP”). The second primary outcome variable was the rate of any VTE associated with TCP (“VTE+TCP”). The secondary outcomes were the rates of the following: any VTE, CVST, MesT, PVT, and TCP. For the secondary outcome variable “Any VTE”, we counted the number of patients, and for all other variables we counted the number of events. See Appendix I in the Supplementary material for a complete list of the considered diagnoses and their ICD-10 codes.

### Study cohorts

The first vaccine cohort consisted of all people 10 years-old and over who received the first dose of Comirnaty®, Spikevax®, or VaxZevria® from 1 January 2021 to 18 April 2021 (week 16). We excluded vaccine recipients who were diagnosed of Covid-19 in the 21 days following vaccination with the first or with the second dose, and we also excluded people who were diagnosed of Covid-19 and received a first or a second vaccine dose in the following 21 days. The second vaccine cohort consisted of all people who received the second dose of Comirnaty® or of Spikevax® up to April 18, with the same exclusion criteria.

The Covid-19 cohort consisted of all people 10 years-old and over who were diagnosed of Covid-19 from 1 January to 18 April 2021. The reference population for the vaccinated cohorts and the Covid-19 cohort was the population on 1 January 2021.

The general population in Catalonia on 1 January 2019 with follow up to 31 December 2019 served as prespecified comparator cohort (2020 was discarded as a reference because in the first months of the pandemic a sharp increase of the incidence of VTE was recorded, apparently related to Covid-19; figure S1 in the Supplementary material). 21-day incidence rates were calculated by assuming a uniform distribution of the outcomes of interest throughout the year.

The study dataset consisted of the data collected and registered in the CMBD up to 9 July 2021.

### Statistical analyses

We calculated the incidence rates of each variable as the number of cases per 100 000 in the 21 days following vaccination or following the day of the positive test, in the total population of each cohort, and by age and sex. We calculated the age- and sex-standardised incidence rates and differences per 100 000, and standardised rate ratios (SRR), using the background cohort as the reference population. This direct standardisation approach is particularly appropriate to compare estimates across cohorts with different age and sex distributions. We obtained 95% CIs from Poisson distribution for the crude estimates, and from gamma, normal and lognormal distributions for standardised incidences, differences, and rate ratios, respectively. Stratified analyses were performed by sex and by age groups (by decades and by larger groups, i.e., 10–39, 40–69, 70–89, and >89 years) for the outcome variables with highest number of cases.

FF, EC, JRL, and XV carried out the data analysis. All analyses were performed with SAS® 9.4 (SAS Institute Inc., Cary, NC, USA).

The study was approved by the Research Ethics Committee of HU Vall d’Hebron, Institut Català de la Salut, Barcelona (protocol reference number EOM(AG)051/2021(5870).

The <u>study protocol</u> and its <u>Methods section translated to English</u> are available at the FICF website.

## Results

### Venous thromboembolism and thrombocytopenia in the reference population

The annual background incidence of unusual site VTE or VTE associated with TCP was 37.34 per 100 000, which corresponds to 2.15 per 100 000 during a 21 day period. The incidence of VTE+TCP was 6.53 per 100 000, which corresponds to 0.38 per 100 000 during a 21 day period. The background incidence of all the primary and secondary outcome variables increased with age, and it was higher in men than in women, in particular portal vein thrombosis (tables S1 and S2).

### Study cohorts

Between 1 January 2021 and 18 April 2021, 1 672 110 people had received the first dose of the vaccine; 9 391 were excluded because they were diagnosed of Covid-19 in the 21 days before or following the first dose, leaving 1 662 719 people in the first dose study cohort (tables S3 and S4). 625 790 people had received the second dose; 3 012 were excluded because they were diagnosed of Covid-19 in the 21 days before or after the second dose, leaving 622 778 people in the second dose study cohort (tables S5 and S6).

In the same period, 200 210 people were diagnosed of Covid-19. Of these, 9 594 were excluded because they received a first or a second dose of a vaccine in the 21 days before or following the diagnosis, leaving 190 616 people in the Covid-19 cohort (tables S7 and S8).

The study cohort vaccinated with the first dose included 989 118 people vaccinated with Comirnaty®, 83 009 with Spikevax®, and 590 137 with VaxZevria®. More than 70 percent of the doses of Comirnaty® were used for people over 70 years of age, while 56 percent of the doses of Spikevax® were used for people 40 to 69 years-old. All doses of VaxZevria® were used for people younger than 70 years old (table S9).

### Venous thromboembolism and thrombocytopenia in the vaccinated cohorts following the first and the second dose

In the cohort vaccinated with the first dose of any Covid-19 vaccine, the 21 day incidence rate of unusual site VTE or VTE associated with TCP was 5.65 per 100 000 doses (94 cases, standardised difference, 2.53 per 100 000 doses [95% CI, 1.04 to 4.00], standardised rate ratio [SRR], 2.17 [95% CI 1.58 to 2.98]).

In the cohort vaccinated with the second dose, the 21 day incidence of unusual site VTE or VTE associated with TCP was 7.23 per 100 000 (45 cases, standardised difference, 4.07 per 100 000 doses [95% CI, 1.43 to 6.70], SRR, 2.89 (95% CI 1.89 to 4.41]), and the incidence of VTE+TCP was 0.80 per 100 000 (five cases, standardised difference, 0.92 [95% CI, −0.45 to 2.30], SRR, 3.45 [95% CI 1.19 to 9.98]) (table 1).

**Table 1.** **Number of cases, standardised incidence and standardised rate difference of the outcome variables: background (2019), in vaccinated people following the first and following the second dose, and in patients with Covid−19. Incidence is expressed as number per 100 000 in 21 days (95% confidence interval).**

|  | Population,<br>N | Unusual<br>thrombosis and<br>TCP<sup>a</sup> | VTE+TCP<sup>b</sup> | Any VTE<sup>c</sup> | TCP<sup>d</sup> |
| --- | --- | --- | --- | --- | --- |
| **Background (2019)** |  |  |  |  |  |
| N | 7 013 040 | 2 619 | 458 | 9 539 | 7 368 |
| Annual incidence, n/100 000 |  | 37.34<br>(35.94–38.80) | 6.53<br>(5.96–7.16) | 136.02<br>(133.32–138.78) | 105.06<br>(102.69–107.49) |
| 21 day incidence, n/100 000 |  | 2.15<br>(2.07–2.24) | 0.38<br>(0.34–0.41) | 7.85<br>(7.69–8.01) | 6.06<br>(5.92–6.20) |
| **Vaccinated first dose** |  |  |  |  |  |
| N | 1 662 719 | 94 | 14 | 345 | 265 |
| 21 day incidence, n/100 000 |  | 5.65<br>(4.62–6.92) | 0.84<br>(0.50–1.42) | 20.75<br>(18.67–23.06) | 15.94<br>(14.13–17.98) |
| Excess rate, n/100 000 |  | 3.50<br>(2.35–4.64) | 0.46<br>(0.02–0.91) | 12.90<br>(10.71–15.10) | 9.88<br>(7.95–11.80) |
| Standardised 21 day incidence,<br>n/100 000 |  | 4.68<br>(3.32–6.40) | 0.93<br>(0.36–1.94) | 13.11<br>(10.82–15.76) | 13.14<br>(10.37–16.35) |
| Standardised 21 day excess rate,<br>n/100 000 |  | 2.53<br>(1.04–4.00) | 0.55<br>(–0.16–1.26) | 5.26<br>(2.85–7.68) | 7.08<br>(4.15–10.01) |
| **Vaccinated second dose** |  |  |  |  |  |
| N | 622 778 | 45 | 5 | 160 | 133 |
| 21 day incidence, n/100 000 |  | 7.23<br>(5.39–9.68) | 0.80<br>(0.33–1.93) | 25.69<br>(22.00–30.00) | 21.36<br>(18.02–25.31) |
| Excess rate, n/100 000 |  | 5.07<br>(2.96–7.18) | 0.43<br>(–0.28–1.13) | 17.84<br>(13.86–21.83) | 15.29<br>(11.66–18.93) |
| Standardised 21 day incidence,<br>n/100 000 |  | 6.22<br>(3.87–9.49) | 1.30<br>(0.31–3.46) | 15.53<br>(11.81–20.08) | 16.68<br>(12.51–21.78) |
| Standardised 21 day excess rate,<br>n/100 000 |  | 4.07<br>(1.43–6.70) | 0.92<br>(–0.45–2.30) | 7.68<br>(3.71–11.65) | 10.62<br>(6.14–15.09) |
| **Covid-19** |  |  |  |  |  |
| N | 190 616 | 62 | 33 | 686 | 442 |
| 21 day incidence, n/100 000 |  | 32.53<br>(25.36–41.72) | 17.31<br>(12.31–24.35) | 359.89<br>(333.94–387.85) | 231.88<br>(211.24–254.54) |
| Excess rate, n/100 000 |  | 30.37<br>(22.27–38.47) | 16.94<br>(11.03–22.84) | 352.04<br>(325.11–378.97) | 225.82<br>(204.20–247.44) |
| Standardised 21 day incidence,<br>n/100 000 |  | 37.76<br>(28.90–48.41) | 19.97<br>(13.71–28.04) | 413.42<br>(382.80–445.78) | 262.93<br>(238.77–288.83) |
| Standardised 21 day excess rate,<br>n/100 000 |  | 35.60<br>(26.15–45.06) | 19.59<br>(12.74–26.45) | 405.57<br>(374.37–436.77) | 256.87<br>(232.13–281.62) |

<sup>a</sup> Any of the following: cerebral venous sinus thrombosis, mesenteric thrombosis, portal vein thrombosis, or any venous thromboembolism associated with thrombocytopenia.
<sup>b</sup> Patients with a simultaneous diagnosis of venous thromboembolism and thrombocytopenia.
<sup>c</sup> Any diagnosis of venous thromboembolism.
<sup>d</sup> Thrombocytopenia (idiopathic, secondary, not specified).

We observed increased excess rates of any VTE (5.26 per 100 000 [95% CI, 2.85 to 7.68]) and of thrombocytopenia (7.08 per 100 000 [95% CI, 4.15 to 10.01]) in the cohort vaccinated with the first dose. Following the second dose, the excess rate of any VTE was 7.68 cases per 100 000 doses [95% CI, 3.71 to 11.65], and the excess rate of thrombocytopenia was 10.62 per 100 000 doses [95% CI, 6.14 to 15.09] (table 1). The incidence rates of all the outcome variables following the first dose of Comirnaty® and Spikevax® were not different from those following the second dose (tables S4 and S6). The risk following the second dose of VaxZevria® could not be evaluated, because only 178 individuals had received it when its use was temporarily halted and deferred from 12 to 16 weeks after the first dose at the end of March.

### Venous thromboembolism and thrombocytopenia in the Covid-19 cohort

In the Covid-19 cohort, the 21 day event rate of unusual site VTE or VTE associated with TCP was 32.53 per 100 000 patients (standardised difference, 35.60 per 100 000 [95% CI, 26.15 to 45.06], SRR, 17.52 [95% CI, 13.60 to 22.58]) (table 1). The 21 day event rate of VTE+TCP was 17.31 per 100 000 (standardised difference, 19.59 per 100 000 patients [95% CI, 12.74 to 26.45], SRR, 53.01 [95% CI 37.16 to 75.62])).

### Sex and age

Both in the vaccinated cohorts and in the Covid-19 cohort, the rates of all the outcome variables in men were approximately two-fold those in women (tables S4, S6 and S8). The excess rate estimates of “Any venous thromboembolism” were also higher in men than in women in all the age strata. Excess rate estimates increased by age in the Covid-19 cohort, but not in the vaccinated cohorts (figure 1 and tables S10 and S11).

**Figure 1.**
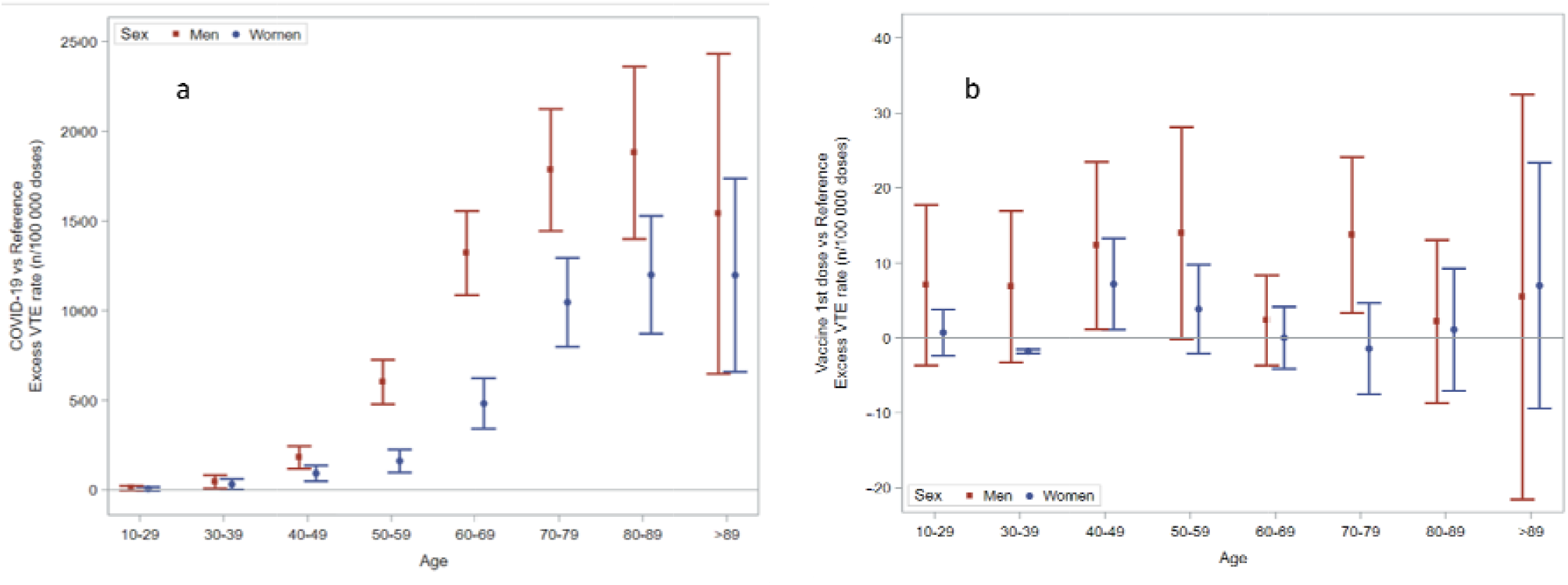
Excess rate of any venous thromboembolism by age and sex groups, in the Covid-19 cohort (a) and following the first dose of Covid-19 vaccines (b). The scale of the ordinal axis differs by 50-fold.

### Pattern of specific diagnoses of venous thromboembolism and thrombocytopenia in the study cohorts

Of 9 539 cases of venous thromboembolism in the reference population, 2 619 (27.5%) were in unusual anatomical sites (table S1). This proportion was similar in the two vaccinated cohorts (94 of 345 [27.3 percent] following the first dose, and 45 of 160 [28.1 percent] following the second dose, tables S3 and S5), but it was significantly lower in the Covid-19 cohort (62 of 686, 9.0 percent], table S7and table S12).

### Event rates with the different vaccines

Ninety-four vaccine recipients developed unusual site VTE or VTE associated with TCP in the 21 days following the first vaccine dose, 58 after Comirnaty® (4.06 per 100 000 [95% CI, 2.12 to 7.02]), 19 after Spikevax® (18.84 per 100 000 doses [95% CI, 10.50 to 31.76), and 17 after VaxZevria® (2.01 [95% CI, 0.99 to 4.05]).

Fourteen people developed VTE+TCP following the first dose of a Covid-19 vaccine (six Comirnaty®, three Spikevax®, and five VaxZevria®), and five following the second dose (four Comirnaty®, one Spikevax®, tables S12 and S13).

The rate of any venous thromboembolism was also significantly higher with Spikevax® (241 cases, 71.67 per 100 000 doses [95% CI, 52.47 to 95.62]) than with Comirnaty® (68 cases, 10.98 per 100 000 [95% CI, 8.05 to 14.72]) and it was lower with VaxZevria® (36 cases, 3.68 [95% CI, 2.27 to 6.01]) after the first dose and after the second dose (table 2).

**Table 2.** **Standardised incidence (95% confidence interval) of the outcomes of interest in the 21 days following the first and the second dose of each vaccine (n/100 000 doses).**

|  | Doses, n | Unusual thrombosis and TCP<sup>a</sup> | VTE+TCP<sup>b</sup> | Any VTE<sup>c</sup> | CVST<sup>d</sup> | MesT<sup>e</sup> | PVT<sup>f</sup> | TCP<sup>g</sup> |
| --- | --- | --- | --- | --- | --- | --- | --- | --- |
| **First dose** |  |  |  |  |  |  |  |  |
| Comirnaty® | 989 118 | 4.06<br>(2.12–7.02) | 0.73<br>(0.05–2.85) | 10.98<br>(8.05–14.72) | – | 0.79<br>(0.22–2.38) | 2.30<br>(0.93–4.75) | 10.51<br>(6.87–15.23) |
| Spikevax® | 83 009 | 18.84<br>(10.50–31.76) | 2.45<br>(0.46–9.63) | 71.67<br>(52.47–95.62) | – | 3.66<br>(0.73–11.89) | 12.73<br>(5.94–24.36) | 81.62<br>(60.35–107.82) |
| VaxZevria® | 590 137 | 2.01<br>(0.99–4.05) | 0.74<br>(0.17–2.55) | 3.68<br>(2.27–6.01) | 0.42<br>(0.09–2.01) | 0.21<br>(0.04–1.74) | 0.63<br>(0.12–2.39) | 3.02<br>(1.59–5.48) |
| **Any** | **1 662 719** | **4.68**<br>**(3.32–6.40)** | **0.93**<br>**(0.36–1.94)** | **13.11**<br>**(10.82–15.76)** | **0.29**<br>**(0.08–0.90)** | **0.97**<br>**(0.49–1.80)** | **2.49**<br>**(1.49–3.90)** | **13.14**<br>**(10.37–16.35)** |
| **Second dose** |  |  |  |  |  |  |  |  |
| Comirnaty® | 585 931 | 5.31<br>(2.99–8.72) | 1.27<br>(0.23–3.70) | 10.72<br>(7.54–14.85) | 0.02<br>(0.00–1.50) | 1.68<br>(0.70–3.69) | 2.34<br>(0.88–4.99) | 11.67<br>(8.31–16.01) |
| Spikevax® | 36 669 | 17.37<br>(6.79–38.21) | 3.15<br>(0.08–18.89) | 54.97<br>(34.29–84.91) | – | 4.58<br>(0.52–20.19) | 9.64<br>(2.51–27.55) | 61.70<br>(36.37–97.81) |
| **Any** | **622 600** | **6.22**<br>**(3.87–9.49)** | **1.30**<br>**(0.31–3.46)** | **15.53**<br>**(11.81–20.08)** | **0.02**<br>**(0.00–1.32)** | **1.81**<br>**(0.82–3.69)** | **3.09**<br>**(1.49–5.68)** | **16.68**<br>**(12.51–21.78)** |

<sup>a</sup> Any of the following: cerebral venous sinus thrombosis, mesenteric thrombosis, portal vein thrombosis, or any VTE associated with thrombocytopenia.
<sup>b</sup> Patients with simultaneous diagnoses of venous thromboembolism and thrombocytopenia.
<sup>c</sup> Any diagnosis of venous thromboembolism.
<sup>d</sup> Cerebral venous sinus thrombosis.
<sup>e</sup> Mesenteric thrombosis.
<sup>f</sup> Portal vein thrombosis.
<sup>g</sup> Thrombocytopenia (idiopathic, secondary, not specified).

### Case fatality

Among 139 vaccine recipients who developed unusual site VTE or VTE associated with TCP following vaccination, 25 died (18 percent; 15 following the first dose, 10 following the second dose). Of 19 vaccine recipients who developed VTE+TCP in the 21 days following vaccination, three died (15.8 percent; two following the first dose, one following the second dose).

## Discussion

In this population based study we observed slightly increased rates of venous thromboembolism in usual and in unusual anatomical sites, and increased rates of venous thromboembolism associated with thrombocytopenia, following vaccination with both mRNA-based and adenovirus vector-based Covid-19 vaccines. We found an excess event rate of unusual site venous thrombosis and of thrombosis associated with thrombocytopenia of 2.5 cases per 100 000 following the first dose, and four per 100 000 following the second dose. The co-primary variable of any venous thromboembolism associated with thrombocytopenia showed a similar trend, but the number of exposed cases was low, and the difference with respect to the reference population was not statistically significant. In the vaccinated cohorts, we found excess rates of any venous thromboembolism of 5.3 and 7.7 cases per 100 000 doses, following the first and the second dose, respectively. The event rate of thrombocytopenia was also increased, by 7.1 and by 10.6 cases per 100 000, respectively.

The excess rates in the vaccinated cohorts were many-fold lower than those in the contemporary cohort of incident cases of Covid-19.

In the reference population, the rates of both usual and unusual site venous thromboembolism and of thrombocytopenia increased with age, and they were two-fold in men compared with women. In the vaccinated cohorts, the rates and the excess rates of all outcome variables were two-fold in men compared with women, but they did not show any particular increase in any age group.

Unexpectedly, the rate of any venous thromboembolism and the rate of thrombocytopenia following the first dose of VaxZevria® were lower than following the first dose of Comirnaty® and the first dose of Spikevax®. Also unexpectedly, the incidence rates of all outcomes except cerebral venous sinus thrombosis were higher with Spikevax® than with Comirnaty® or with VaxZevria®. During the vaccination campaign, Spikevax® and Comirnaty® were targeted to health professionals, but also to nursing home residents, those over 80 years of age, and other high risk groups. Our analyses were adjusted for age and sex, but not for other risk factors, and therefore we cannot conclude on any differences of risk between vaccines, except that mRNA-based Covid-19 vaccines are associated with an increased rate of venous thrombosis, thrombocytopenia, or both, which is of the same order of magnitude as the rate associated with adenovirus vector-based Covid-19 vaccines.

We did not observe a higher rate of the outcomes of interest following the second dose of Comirnaty® or of Spikevax®, compared to their first doses. Although we cannot exclude possible selection bias (e.g., by depletion of susceptible individuals, i.e., people who had a bad experience following the first dose would be less likely to present for the second shot), the risk of venous thromboembolism following the second dose was of the same order of magnitude as following the first dose. The risk following the second dose of VaxZevria® could not be evaluated, because on March 30 the administration of the second dose was deferred from 12 to 16 weeks after the first dose.

The present study is the first which has examined the incidence of various diagnostic definitions of venous thromboembolism and thrombocytopenia by age and by sex, in contemporary cohorts of people vaccinated with the first or the second dose of various Covid-19 vaccines and people with Covid-19, compared with the background incidence in the same source population and dataset.

Covid-19 infection is associated with thrombotic complications, whose complete pathophysiology is not well understood. Cerebral venous sinus thrombosis,^10,11^ mesenteric thrombosis,^12^ and portal vein thrombosis^13,14^ may be presenting features or may occur at a late stage in the disease. Thrombocytopenia can also be a complication of Covid-19 disease.^15,16^ Thrombocytopenia and venous thromboembolism may be triggered by platelet activating antibodies against platelet factor 4, resulting in blood clotting and consumption of platelets. These antibodies have been identified in patients with Covid-19^17^ and in people vaccinated with VaxZevria® who experienced the clots.^18,19^ It is therefore logical to suspect that the immune responses associated with Covid-19 infection and Covid-19 vaccination may share some similarities that would increase the risk of thrombotic events in susceptible individuals.

The incidence of venous thromboembolism and of thrombocytopenia was one order of magnitude higher in the Covid-19 cohort, compared with the reference population and with the vaccinated cohorts. However, in the reference population and in the vaccinated cohorts the proportions of specific diagnoses of unusual site venous thrombosis were between 25 and 30 percent, while in the Covid-19 cohort it was 8.5 percent. Thrombosis associated with Covid-19 is a research challenge which should be addressed through international collaboration of specialists in immunology, virology, haematology and others, with a primary focus on Covid-19 patients, rather than on Covid-19 vaccines.

In summary, our data indicate that vaccination against Covid-19 increases the rate of venous thromboembolism in usual and in unusual anatomic locations, with or without thrombocytopenia. Contrary to the initial suggestions from spontaneous reporting, this risk is shared by adenovirus vector-based and mRNA-based Covid-19 vaccines, it is higher in men than in women, and it does not concentrate in any particular age group.

Spontaneous reporting of suspicions of adverse effects of medicines or vaccines has uncovered hundreds of signals of possible adverse effects of medicines and vaccines, as the experience with Covid-19 vaccines and thrombosis has shown. However, uncommon and unexpected clinical conditions, e.g., cerebral venous sinus thrombosis in previously healthy young women, are powerful triggers of reporting, while common conditions, e.g., thrombosis in those over 70, do not draw the same attention and therefore they are less likely to be reported.

### Strengths and weaknesses of this study

Our study has several strengths. First, it was population-based and all potentially vaccinated age groups were included. This allowed the analysis of rates by age groups.

Second, we compared the incidence of the outcomes of interest in vaccinated people with the background incidence, and we also examined a contemporary Covid-19 cohort originated from the same population and in the same dataset. This avoids potential systematic error and confounding caused by heterogeneity between healthcare databases.^20^

Third, we examined the excess rates of various diagnostic definitions of usual and unusual site venous thromboembolism, with or without thrombocytopenia. This allows the analysis of the rates of specific clinical outcomes, and at the same time it gives an overview of the epidemiology of venous thromboembolism in people with Covid-19 and following vaccination.

Fourth, rather than examining the risks associated to a particular vaccine, we included all Covid-19 vaccines in use in our country.

The yearly incidence rates of the outcome variables in the reference population and the case-fatality rates were similar to those found in other recent studies.^21-24^

Our findings should however be interpreted in the context of their limitations.

First, we did not review the clinical records and haematological data of patients with venous thromboembolism or thrombocytopenia, and therefore we were not able to evaluate laboratory and clinical data which could help to clarify whether the mechanism of thrombosis and thrombocytopenia after vaccination shares some features with those observed in Covid-19 patients.

Second, we did not exclude individuals with a history of venous thromboembolism in the 12 months before. This may have introduced some degree of bias and confounding.

Third, the data are entered into the CMBD database only after patient discharge or patient death, but some centres take a few days to complete registration. Therefore, a small proportion of patients with a hospital stay of more than four weeks may have been missed. The recruitment of the cohorts was done up to 18 April, and the time window of observation was 21 days (i.e., up to 9 May). We closed the dataset on July 9, so that some patients with a long hospital stay of more than six to eight weeks may have been missed. This may have underestimated the rates of the outcome variables in both the vaccinated cohorts and the Covid-19 cohort.

Fourth, although we estimated excess rates by age and sex, we did not have access to the clinical records and we could not take into account other risk factors, in particular chronic conditions associated to inflammation, immunosupression, and other. Therefore, our data should be interpreted as evidence that the risk is shared by all three Covid-19 vaccines, although we cannot exclude any qualitative difference in the pattern of venous thromboembolism and thrombocytopenia between the various available vaccines.

Our data refer to only a small ingredient of the global balance of benefits and risks of Covid-19 vaccines. This will of course depend on how effective the available vaccines will be against new emerging variants of SARS-CoV-2, the risk of other possible adverse effects of the vaccines, the readiness of health systems to vaccinate, and social acceptance of vaccination against Covid-19.

## Conclusion

In conclusion, we observed an excess rate of venous thromboembolism and thrombocytopenia following Covid-19 vaccination, of the order of two to four additional cases of unusual site venous thrombosis and eight to ten additional cases of any venous thromboembolism or thrombocytopenia per 100 000 doses. The excess event rates of venous thrombosis and thrombocytopenia were not especially increased in any age group, and they were two-fold in men compared with women. They were higher in recipients of the Spikevax® Covid-19 vaccine, compared with Comirnaty® and lower with VaxZevria®, although we cannot exclude that this was partly due to different vulnerabilities of the populations targeted for with each vaccine. In Covid-19 patients, the rate of venous thromboembolism and thrombocytopenia was many-fold higher, it increased with age, and it was also two-fold in men than in women. Covid-19 and Covid-19 vaccines may share immunological mechanisms leading to thrombosis in a minority of susceptible people.

## Data Availability

Owing to Spanish data protection regulations, individual patients data can be used by health authorities and public health institutions in situations of exceptional relevance for public health. However, these data cannot be shared with third parties. The CMDB provided the results of the analyses as anonimised .xls files, which were archived. Detailed aggregated data are provided in the Supplementary Material of this article. All aggregated data will be available upon reasonable request to the corresponding autor after peer reviewed publication.

https://www.icf.uab.cat/es/download/enllac/assets/pdf/Research%20Protocol%20-%20Covid-19%20vaccines%20and%20thromboembolism%20and%20thrombocytopenia.pdf

https://www.icf.uab.cat/es/download/enllac/assets/pdf/Study%20Protocol%20English%20Translation%20of%20Methods.pdf

## Acknowledgements

On April 15, Professor Bernard Bégaud, Emeritus Professor of Clinical Pharmacology at the University of Bordeaux, gave a scientific seminar on causality and chance in pharmacovigilance signals of Covid-19 vaccines at the Italian Medicines Agency (AIFA). His reflections and the debate at this meeting triggered the analysis reported in ths article. We are also grateful to Dr Gianni Tognoni and Dr Valerio Reggi for their useful comments, and to Dr. Carme Cabezas, Public Health Director General at the Department of Health, Generalitat de Catalunya.

## Notes

**Conflict of interests statement** All authors have completed the ICMJE uniform disclosure form at www.icmje.org/coi_disclosure.pdf and declare: no support from any organisation for the submitted work; no financial relationships with any organisations that might have an interest in the submitted work in the previous three years; no other relationships or activities that could appear to have influenced the submitted work.

### Competing Interest Statement

The authors have declared no competing interest.

### Clinical Trial

For reasons of public health emergency, the study protocol was deposited in the registry of the Ethics Committee which evaluated and approved the protocol. The protocol was published at the institutional web of the Catalan Institute of Pharmacology: https://www.icf.uab.cat/es/download/enllac/assets/pdf/Research%20Protocol%20-%20Covid-19%20vaccines%20and%20thromboembolism%20and%20thrombocytopenia.pdf

### Funding Statement

There was no direct funding of this study. The institutions of the Department of Health where the authors work were not involved in the study design; in the analysis, and interpretation of data; in the writing of the report; or in the decision to submit the paper for publication. All authors had full access to all of the data (including statistical reports and tables) in the study and can take responsibility for the integrity of the data and the accuracy of the data analysis.

### Author Declarations

The study was approved by the Research Ethics Committee of HU Vall Hebron, Institut Catala de la Salut, Barcelona (protocol reference number EOM(AG)051/2021(5870).

### Summary of Updates

Spelling errors have been corrected in the Summary (5th line, year 2019), missing axis labels have been added to figure S1. Details in calculation of background incidence have been added to the Methods section. 172 recipients of a second dose of VaxZevria have been deleted from tables 2 and S13. A sentence in the Discussion regarding the risk of mesenteric thrombosis with Spikevax has been corrected. A spelling error on the number of cases of unusual site VTE or VTE+TCP in the Results section has been corrected.

