## Supplementary Material for "Vaccines against Covid-19, venous thromboembolism, and thrombocytopenia. A population-based retrospective cohort study"

**Appendix I**

**Diagnostic codes**

The following ICD-10 diagnoses were included in the outcome definition Venous thromboembolism:

**Cerebral venous sinus thrombosis**

G08	Intracranial and intraspinal phlebitis and thrombophlebitis

I676	Nonpyogenic thrombosis of intracranial venous system

I636	Cerebral infarction due to cerebral venous thrombosis, nonpyogenic

**Mesenteric thrombosis**

K55011	Focal (segmental) acute (reversible) ischemia of small intestine

K55012	Diffuse acute (reversible) ischemia of small intestine

K55019	Acute (reversible) ischemia of small intestine, extent unspecified

K55021	Focal (segmental) acute infarction of small intestine

K55022	Diffuse acute infarction of small intestine

K55029	Acute infarction of small intestine, extent unspecified

K55031	Focal (segmental) acute (reversible) ischemia of large intestine

K55032	Diffuse acute (reversible) ischemia of large intestine

K55039	Acute (reversible) ischemia of large intestine, extent unspecified

K55041	Focal (segmental) acute infarction of large intestine

K55042	Diffuse acute infarction of large intestine

K55049	Acute infarction of large intestine, extent unspecified

K55051	Focal (segmental) acute (reversible) ischemia of intestine, part unspecified

K55052	Diffuse acute (reversible) ischemia of intestine, part unspecified

K55059	Acute (reversible) ischemia of intestine, part and extent unspecified

K55061	Focal (segmental) acute infarction of intestine, part unspecified

K55062	Diffuse acute infarction of intestine, part unspecified

K55069	Acute infarction of intestine, part and extent unspecified

**Portal vein thrombosis**

I81	Portal vein thrombosis

**Non-limb venous thrombosis**

I821	Thrombophlebitis migrans

I82210	Acute embolism and thrombosis of superior vena cava

I82220	Acute embolism and thrombosis of inferior vena cava

I82290	Acute embolism and thrombosis of other thoracic veins

I823	Embolism and thrombosis of renal vein

I82B11	Acute embolism and thrombosis of right subclavian vein

I82B12	Acute embolism and thrombosis of left subclavian vein

I82B13	Acute embolism and thrombosis of subclavian vein, bilateral

I82B19	Acute embolism and thrombosis of unspecified subclavian vein

I82C11	Acute embolism and thrombosis of right internal jugular vein

I82C12	Acute embolism and thrombosis of left internal jugular vein

I82C13	Acute embolism and thrombosis of internal jugular vein, bilateral

I82C19	Acute embolism and thrombosis of unspecified internal jugular vein

**Venous thrombosis of extremities**

I82401	Acute embolism and thrombosis of unspecified deep veins of right lower extremity

I82402	Acute embolism and thrombosis of unspecified deep veins of left lower extremity

I82403	Acute embolism and thrombosis of unspecified deep veins of lower extremity, bilateral

I82409	Acute embolism and thrombosis of unspecified deep veins of unspecified lower extremity

I82411	Acute embolism and thrombosis of right femoral vein

I82412	Acute embolism and thrombosis of left femoral vein

I82413	Acute embolism and thrombosis of femoral vein, bilateral

I82419	Acute embolism and thrombosis of unspecified femoral vein

I82421	Acute embolism and thrombosis of right iliac vein

I82422	Acute embolism and thrombosis of left iliac vein

I82423	Acute embolism and thrombosis of iliac vein, bilateral

I82429	Acute embolism and thrombosis of unspecified iliac vein

I82431	Acute embolism and thrombosis of right popliteal vein

I82432	Acute embolism and thrombosis of left popliteal vein

I82433	Acute embolism and thrombosis of popliteal vein, bilateral

I82439	Acute embolism and thrombosis of unspecified popliteal vein

I82441	Acute embolism and thrombosis of right tibial vein

I82442	Acute embolism and thrombosis of left tibial vein

I82443	Acute embolism and thrombosis of tibial vein, bilateral

I82449	Acute embolism and thrombosis of unspecified tibial vein

I82491	Acute embolism and thrombosis of other specified deep vein of right lower extremity

I82492	Acute embolism and thrombosis of other specified deep vein of left lower extremity

I82493	Acute embolism and thrombosis of other specified deep vein of lower extremity, bilateral

I82499	Acute embolism and thrombosis of other specified deep vein of unspecified lower extremity

I824Y1	Acute embolism and thrombosis of unspecified deep veins of right proximal lower extremity

I824Y2	Acute embolism and thrombosis of unspecified deep veins of left proximal lower extremity

I824Y3	Acute embolism and thrombosis of unspecified deep veins of proximal lower extremity, bilateral

I824Y9	Acute embolism and thrombosis of unspecified deep veins of unspecified proximal lower extremity

I824Z1	Acute embolism and thrombosis of unspecified deep veins of right distal lower extremity

I824Z2	Acute embolism and thrombosis of unspecified deep veins of left distal lower extremity

I824Z3	Acute embolism and thrombosis of unspecified deep veins of distal lower extremity, bilateral

I824Z9	Acute embolism and thrombosis of unspecified deep veins of unspecified distal lower extremity

I82601	Acute embolism and thrombosis of unspecified veins of right upper extremity

I82602	Acute embolism and thrombosis of unspecified veins of left upper extremity

I82603	Acute embolism and thrombosis of unspecified veins of upper extremity, bilateral

I82609	Acute embolism and thrombosis of unspecified veins of unspecified upper extremity

I82611	Acute embolism and thrombosis of superficial veins of right upper extremity

I82612	Acute embolism and thrombosis of superficial veins of left upper extremity

I82613	Acute embolism and thrombosis of superficial veins of upper extremity, bilateral

I82619	Acute embolism and thrombosis of superficial veins of unspecified upper extremity

I82621	Acute embolism and thrombosis of deep veins of right upper extremity

I82622	Acute embolism and thrombosis of deep veins of left upper extremity

I82623	Acute embolism and thrombosis of deep veins of upper extremity, bilateral

I82629	Acute embolism and thrombosis of deep veins of unspecified upper extremity

I82811	Embolism and thrombosis of superficial veins of right lower extremity

I82812	Embolism and thrombosis of superficial veins of left lower extremity

I82813	Embolism and thrombosis of superficial veins of lower extremities, bilateral

I82819	Embolism and thrombosis of superficial veins of unspecified lower extremity

I82A11	Acute embolism and thrombosis of right axillary vein

I82A12	Acute embolism and thrombosis of left axillary vein

I82A13	Acute embolism and thrombosis of axillary vein, bilateral

I82A19	Acute embolism and thrombosis of unspecified axillary vein

I8000	Phlebitis and thrombophlebitis of superficial vessels of unspecified lower extremity

I8001	Phlebitis and thrombophlebitis of superficial vessels of right lower extremity

I8002	Phlebitis and thrombophlebitis of superficial vessels of left lower extremity

I8003	Phlebitis and thrombophlebitis of superficial vessels of lower extremities, bilateral

I8010	Phlebitis and thrombophlebitis of unspecified femoral vein

I8011	Phlebitis and thrombophlebitis of right femoral vein

I8012	Phlebitis and thrombophlebitis of left femoral vein

I8013	Phlebitis and thrombophlebitis of femoral vein, bilateral

I80201	Phlebitis and thrombophlebitis of unspecified deep vessels of right lower extremity

I80202	Phlebitis and thrombophlebitis of unspecified deep vessels of left lower extremity

I80203	Phlebitis and thrombophlebitis of unspecified deep vessels of lower extremities, bilateral

I80209	Phlebitis and thrombophlebitis of unspecified deep vessels of unspecified lower extremity

I80211	Phlebitis and thrombophlebitis of right iliac vein

I80212	Phlebitis and thrombophlebitis of left iliac vein

I80213	Phlebitis and thrombophlebitis of iliac vein, bilateral

I80219	Phlebitis and thrombophlebitis of unspecified iliac vein

I80221	Phlebitis and thrombophlebitis of right popliteal vein

I80222	Phlebitis and thrombophlebitis of left popliteal vein

I80223	Phlebitis and thrombophlebitis of popliteal vein, bilateral

I80229	Phlebitis and thrombophlebitis of unspecified popliteal vein

I80231	Phlebitis and thrombophlebitis of right tibial vein

I80232	Phlebitis and thrombophlebitis of left tibial vein

I80233	Phlebitis and thrombophlebitis of tibial vein, bilateral

I80239	Phlebitis and thrombophlebitis of unspecified tibial vein

I80291	Phlebitis and thrombophlebitis of other deep vessels of right lower extremity

I80292	Phlebitis and thrombophlebitis of other deep vessels of left lower extremity

I80293	Phlebitis and thrombophlebitis of other deep vessels of lower extremity, bilateral

I80299	Phlebitis and thrombophlebitis of other deep vessels of unspecified lower extremity

I803	Phlebitis and thrombophlebitis of lower extremities, unspecified

**Nonspecific venous thrombosis**

I82890	Acute embolism and thrombosis of other specified veins

I8290	Acute embolism and thrombosis of unspecified vein

I808	Phlebitis and thrombophlebitis of other sites

I809	Phlebitis and thrombophlebitis of unspecified site

**Pulmonary embolism**

I2601	Septic pulmonary embolism with acute cor pulmonale

I2602	Saddle embolus of pulmonary artery with acute cor pulmonale

I2609	Other pulmonary embolism with acute cor pulmonale

I2690	Septic pulmonary embolism without acute cor pulmonale

I2692	Saddle embolus of pulmonary artery without acute cor pulmonale

I2699	Other pulmonary embolism without acute cor pulmonale

**Thrombocytopenia**

D693	Immune thrombocytopenic purpura

D6959	Other secondary thrombocytopenia

D696	Thrombocytopenia, unspecified

D7582	Heparin induced thrombocytopenia (HIT)

**Appendix II**

**Figure S1.** Weekly number of cases of selected diagnoses of venous thromboembolism in 2020, recorded in the preliminary exploration of the database.

**Table S1.** **Reference population (2019, weeks 1 to 52): number of cases of the selected diagnoses, by age and sex.**

| Age | n | VTE^a^ | CVST^b^ | MesT^c^ | PVT^d^ | TCP^e^ | VTE+TCP^f^ |
| --- | --- | --- | --- | --- | --- | --- | --- |
| 10–19 | 826 219 | 68 | 9 | 1 | 5 | 130 | 6 |
| 20–29 | 847 320 | 160 | 4 | 4 | 9 | 284 | 9 |
| 30–39 | 1 078 419 | 364 | 6 | 20 | 20 | 486 | 13 |
| 40–49 | 1 312 392 | 738 | 20 | 40 | 90 | 534 | 42 |
| 50–59 | 1 072 450 | 1 357 | 15 | 101 | 224 | 918 | 83 |
| 60–69 | 821 399 | 1 883 | 17 | 177 | 273 | 1 291 | 138 |
| 70–79 | 600 679 | 2 366 | 8 | 307 | 252 | 1 778 | 97 |
| 80–89 | 367 307 | 2 097 | 9 | 285 | 143 | 1 554 | 56 |
| >89 | 86 855 | 506 | 1 | 108 | 13 | 393 | 14 |
| **Total** | **7 013 040** | **9 539** | **89** | **1 043** | **1 029** | **7 368** | **458** |
| 10–39 | 2 751 958 | 592 | 19 | 25 | 34 | 900 | 28 |
| 40–69 | 3 206 241 | 3 978 | 52 | 318 | 587 | 2,743 | 263 |
| 70–89 | 967 986 | 4 463 | 17 | 592 | 395 | 3,332 | 153 |
| >89 | 86 855 | 506 | 1 | 108 | 13 | 393 | 14 |
| Sex | | | | | | | |
| Men | **3 426 332** | **5 134** | **51** | **552** | **702** | **4 292** | **301** |
| Women | **3 586 708** | **4 405** | **38** | **491** | **327** | **3 076** | **157** |

^a^ Any diagnosis of venous thromboembolism.

^b^ Cerebral venous sinus thrombosis.

^c^ Mesenteric thrombosis.

^d^ Portal vein thrombosis.

^e^ Thrombocytopenia (idiopathic, secondary, not specified).

^f^ Patients with simultaneous diagnoses of venous thromboembolism and thrombocytopenia.

**Table S2.** **Reference population (2019, weeks 1 to 52): 21 day incidence of the selected diagnoses, by age and sex, n/100 000.**

| Age | n | VTE^a^ | CVST^b^ | MesT^c^ | PVT^d^ | TCP^e^ | VTE+TCP^f^ |
| --- | --- | --- | --- | --- | --- | --- | --- |
| 10–19 | 826 219 | 0.47 | 0.06 | 0.01 | 0.03 | 0.91 | 0.04 |
| 20–29 | 847 320 | 1.09 | 0.03 | 0.03 | 0.06 | 1.93 | 0.06 |
| 30–39 | 1 078 419 | 1.95 | 0.03 | 0.11 | 0.11 | 2.60 | 0.07 |
| 40–49 | 1 312 392 | 3.24 | 0.01 | 0.18 | 0.40 | 2.35 | 0.18 |
| 50–59 | 1 072 450 | 7.30 | 0.08 | 0.54 | 1.20 | 4.94 | 0.45 |
| 60–69 | 821 399 | 13.23 | 0.12 | 1.24 | 1.92 | 9.07 | 0.97 |
| 70–79 | 600 679 | 22.72 | 0.08 | 2.95 | 2.42 | 17.08 | 0.93 |
| 80–89 | 367 307 | 32.94 | 0.14 | 4.48 | 2.25 | 24.41 | 0.88 |
| >89 | 86 855 | 33.61 | 0.07 | 7.17 | 0.86 | 26.10 | 0.93 |
| **Total** | **7 013 040** | **7.85** | **0.07** | **0.86** | **0.85** | **6.06** | **0.38** |
| 10–39 | 2 751 958 | 1.24 | 0.04 | 0.05 | 0.07 | 1.89 | 0.06 |
| 40–69 | 3 206 241 | 7.16 | 0.09 | 0.57 | 1.06 | 4.94 | 0.47 |
| 70–89 | 967 986 | 26.60 | 0.10 | 3.53 | 2.35 | 19.86 | 0.91 |
| >89 | 86 855 | 33.61 | 0.07 | 7.17 | 0.86 | 26.10 | 0.93 |
| Sex | | | | | | | |
| Men | **3 426 332** | **8.64** | **0.09** | **0.93** | **1.18** | **7.23** | **0.51** |
| Women | **3 586 708** | **7.09** | **0.06** | **0.79** | **0.53** | **4.95** | **0.25** |

^a^ Any diagnosis of venous thromboembolism.

^b^ Cerebral venous sinus thrombosis.

^c^ Mesenteric thrombosis.

^d^ Portal vein thrombosis.

^e^ Thrombocytopenia (idiopathic, secondary, not specified).

^f^ Patients with simultaneous diagnoses of venous thromboembolism and thrombocytopenia.

**Table S3.** **People vaccinated with the first dose of a Covid-19 vaccine (2021, weeks 1 to 16): number of cases of the selected diagnoses by age and sex in the 21 days following vaccination.**

| Age | n | VTE^a^ | CVST^b^ | MesT^c^ | PVT^d^ | TCP^e^ | VTE+TCP^f^ |
| --- | --- | --- | --- | --- | --- | --- | --- |
| 10–19 | 6 990 | 1 | 0 | 0 | 0 | 2 | 0 |
| 20–29 | 82 341 | 2 | 0 | 0 | 0 | 6 | 0 |
| 30–39 | 104 149 | 3 | 0 | 0 | 2 | 7 | 1 |
| 40–49 | 151 535 | 18 | 2 | 1 | 2 | 10 | 2 |
| 50–59 | 139 712 | 19 | 0 | 2 | 5 | 20 | 1 |
| 60–69 | 434 153 | 63 | 3 | 3 | 9 | 51 | 4 |
| 70–79 | 337 017 | 94 | 1 | 7 | 18 | 75 | 5 |
| 80–89 | 324 708 | 112 | 0 | 13 | 6 | 69 | 1 |
| >89 | 82 114 | 33 | 0 | 4 | 2 | 25 | 0 |
| **Total** | **1 662 719** | **345** | **6** | **30** | **44** | **265** | **14** |
| 10–39 | 193 480 | 6 | 0 | 0 | 2 | 15 | 1 |
| 40–69 | 725 400 | 100 | 5 | 6 | 16 | 81 | 7 |
| 70–89 | 661 725 | 206 | 1 | 20 | 24 | 144 | 6 |
| >89 | 82 114 | 33 | 0 | 4 | 2 | 25 | 0 |
| Sex | | | | | | | |
| Men | **669 108** | **185** | **2** | **17** | **32** | **163** | **8** |
| Women | **993 611** | **160** | **4** | **13** | **12** | **102** | **6** |

^a^ Any diagnosis of venous thromboembolism.

^b^ Cerebral venous sinus thrombosis.

^c^ Mesenteric thrombosis.

^d^ Portal vein thrombosis.

^e^ Thrombocytopenia (idiopathic, secondary, not specified).

^f^ Patients with simultaneous diagnoses of venous thromboembolism and thrombocytopenia.

**Table S4.** **People vaccinated with the first dose of a Covid-19 vaccine (2021, weeks 1 to 16): 21 day crude incidence of the selected diagnoses by age and sex, n/100 000.**

| Age | n | VTE^a^ | CVST^b^ | MesT^c^ | PVT^d^ | TCP^e^ | VTE+TCP^f^ |
| --- | --- | --- | --- | --- | --- | --- | --- |
| 10–19 | 6 990 | 14.31 | 0 | 0 | 0 | 28.61 | 0 |
| 20–29 | 82 341 | 2.43 | 0 | 0 | 0 | 7.29 | 0 |
| 30–39 | 104 149 | 2.88 | 0 | 0 | 1.92 | 6.72 | 0.96 |
| 40–49 | 151 535 | 11.88 | 1.32 | 0.66 | 1.32 | 6.60 | 1.32 |
| 50–59 | 139 712 | 13.60 | 0 | 1.43 | 3.58 | 14.32 | 0.72 |
| 60–69 | 434 153 | 14.51 | 0.69 | 0.69 | 2.07 | 11.75 | 0.92 |
| 70–79 | 337 017 | 27.89 | 0.30 | 2.08 | 5.34 | 22.25 | 1.48 |
| 80–89 | 324 708 | 34.49 | 0 | 4.00 | 1.85 | 21.25 | 0.31 |
| >89 | 82 114 | 40.19 | 0 | 4.87 | 2.44 | 30.45 | 0 |
| **Total** | **1 662 719** | **20.75** | **0.36** | **1.80** | **2.65** | **15.94** | **0.84** |
| 10–39 | 1934 80 | 3.10 | 0 | 0 | 1.03 | 7.75 | 0.52 |
| 40–69 | 725 400 | 13.79 | 0.69 | 0.83 | 2.21 | 11.17 | 0.96 |
| 70–89 | 661 725 | 31.13 | 0.15 | 3.02 | 3.63 | 21.76 | 0.91 |
| >89 | 82 114 | 40.19 | 0 | 4.87 | 2.44 | 30.45 | 0 |
| Sex | | | | | | | |
| Men | **669 108** | **27.65** | **0.30** | **2.54** | **4.78** | **24.36** | **1.20** |
| Women | **993 611** | **16.10** | **0.40** | **1.31** | **1.21** | **10.27** | **0.60** |

^a^ Any diagnosis of venous thromboembolism.

^b^ Cerebral venous sinus thrombosis.

^c^ Mesenteric thrombosis.

^d^ Portal vein thrombosis.

^e^ Thrombocytopenia (idiopathic, secondary, not specified).

^f^ Patients with simultaneous diagnoses of venous thromboembolism and thrombocytopenia.

**Table S5.** **People vaccinated with the second dose of of Comirnaty® or SpikeVax® (2021, weeks 1 to 16): number of cases of the selected diagnoses by age and sex.**

| Age | n | VTE^a^ | CVST^b^ | MesT^c^ | PVT^d^ | TCP^e^ | VTE+TCP^f^ |
| --- | --- | --- | --- | --- | --- | --- | --- |
| 10–19 | 3 371 | 0 | 0 | 0 | 0 | 0 | 0 |
| 20–29 | 40 536 | 0 | 0 | 0 | 0 | 2 | 0 |
| 30–39 | 46 476 | 3 | 0 | 0 | 1 | 1 | 1 |
| 40–49 | 66 088 | 4 | 0 | 0 | 1 | 6 | 0 |
| 50–59 | 72 243 | 12 | 0 | 1 | 3 | 5 | 0 |
| 60–69 | 51 907 | 15 | 0 | 2 | 2 | 7 | 1 |
| 70–79 | 40 390 | 15 | 0 | 4 | 3 | 34 | 2 |
| 80–89 | 229 416 | 85 | 1 | 9 | 10 | 52 | 1 |
| >89 | 72 351 | 26 | 0 | 2 | 1 | 26 | 0 |
| **Total** | **622 778** | **160** | **1** | **18** | **21** | **133** | **5** |
| 10–39 | 90 383 | 3 | 0 | 0 | 1 | 3 | 1 |
| 40–69 | 190 238 | 31 | 0 | 3 | 6 | 18 | 1 |
| 70–89 | 269 806 | 100 | 1 | 13 | 13 | 86 | 3 |
| >89 | 72 351 | 26 | 0 | 2 | 1 | 26 | 0 |
| Sex | | | | | | | |
| Men | **210 675** | **73** | **0** | **6** | **14** | **80** | **3** |
| Women | **412 103** | **87** | **1** | **12** | **7** | **53** | **2** |

^a^ Any diagnosis of venous thromboembolism.

^b^ Cerebral venous sinus thrombosis.

^c^ Mesenteric thrombosis.

^d^ Portal vein thrombosis.

^e^ Thrombocytopenia (idiopathic, secondary, not specified).

^f^ Patients with simultaneous diagnoses of venous thromboembolism and thrombocytopenia.

**Table S6.** **People vaccinated with the second dose of Comirnaty® or SpikeVax® (2021, weeks 1 to 16): 21 day crude incidence of the selected diagnoses by age and sex, n/100 000.**

| Age | n | VTE^a^ | CVST^b^ | MesT^c^ | PVT^d^ | TCP^e^ | VTE+TCP^f^ |
| --- | --- | --- | --- | --- | --- | --- | --- |
| 10–19 | 3 371 | 0 | 0 | 0 | 0 | 0 | 0 |
| 20–29 | 40 536 | 0 | 0 | 0 | 0 | 4.93 | 0 |
| 30–39 | 46 476 | 6.45 | 0 | 0 | 2.15 | 2.15 | 2.15 |
| 40–49 | 66 088 | 6.05 | 0 | 0 | 1.51 | 9.08 | 0 |
| 50–59 | 72 243 | 16.61 | 0 | 1.38 | 4.15 | 6.92 | 0 |
| 60–69 | 51 907 | 28.90 | 0 | 3.85 | 3.85 | 13.49 | 1.93 |
| 70–79 | 40 390 | 37.14 | 0 | 9.90 | 7.43 | 84.18 | 4.95 |
| 80–89 | 229 416 | 37.05 | 0.44 | 3.92 | 4.36 | 22.67 | 0.44 |
| >89 | 72 351 | 35.94 | 0 | 2.76 | 1.38 | 35.94 | 0 |
| **Total** | **622 778** | **25.69** | **0.16** | **2.89** | **3.37** | **21.36** | **0.80** |
| 10–39 | 90 383 | 3.32 | 0 | 0 | 1.11 | 3.32 | 1.11 |
| 40–69 | 190 238 | 16.30 | 0 | 1.58 | 3.15 | 9.46 | 0.53 |
| 70–89 | 269 806 | 37.06 | 0.37 | 4.82 | 4.82 | 31.87 | 1.11 |
| >89 | 72 351 | 35.94 | 0 | 2.76 | 1.38 | 35.94 | 0 |
| Sex | | | | | | | |
| Men | **210 675** | **34.65** | **0** | **2.85** | **6.65** | **37.97** | **1.42** |
| Women | **412 103** | **21.11** | **0.24** | **2.91** | **1.70** | **12.86** | **0.49** |

^a^ Any diagnosis of venous thromboembolism.

^b^ Cerebral venous sinus thrombosis.

^c^ Mesenteric thrombosis.

^d^ Portal vein thrombosis.

^e^ Thrombocytopenia (idiopathic, secondary, not specified).

^f^ Patients with simultaneous diagnoses of venous thromboembolism and thrombocytopenia.

**Table S7.** **Incident cases of Covid-19 (2021, weeks 1 to 16): number of cases of the selected diagnoses, by age and sex in the 21 days following diagnosis.**

| Age | n | VTE^a^ | CVST^b^ | MesT^c^ | PVT^d^ | TCP^e^ | VTE+TCP^f^ |
| --- | --- | --- | --- | --- | --- | --- | --- |
| 10–19 | 29 088 | 0 | 0 | 0 | 0 | 3 | 0 |
| 20–29 | 24 660 | 5 | 0 | 0 | 0 | 7 | 0 |
| 30–39 | 27 570 | 11 | 0 | 0 | 0 | 15 | 1 |
| 40–49 | 36 735 | 51 | 0 | 2 | 0 | 42 | 1 |
| 50–59 | 31 020 | 120 | 0 | 0 | 4 | 73 | 6 |
| 60–69 | 18 901 | 172 | 2 | 2 | 6 | 90 | 10 |
| 70–79 | 12 683 | 180 | 0 | 6 | 2 | 122 | 14 |
| 80–89 | 7 572 | 115 | 0 | 3 | 1 | 66 | 1 |
| >89 | 2 387 | 32 | 0 | 0 | 1 | 24 | 0 |
| **Total** | **190 616** | **686** | **2** | **13** | **14** | **442** | **33** |
| 10–39 | 81 318 | 16 | 0 | 0 | 0 | 25 | 1 |
| 40–69 | 86 656 | 343 | 2 | 4 | 10 | 205 | 17 |
| 70–89 | 20 255 | 295 | 0 | 9 | 3 | 188 | 15 |
| >89 | 2 387 | 32 | 0 | 0 | 1 | 24 | 0 |
| Sex | | | | | | | |
| Men | **92 072** | **443** | **1** | **8** | **10** | **292** | **26** |
| Women | **98 544** | **243** | **1** | **5** | **4** | **150** | **7** |

^a^ Any diagnosis of venous thromboembolism.

^b^ Cerebral venous sinus thrombosis.

^c^ Mesenteric thrombosis.

^d^ Portal vein thrombosis.

^e^ Thrombocytopenia (idiopathic, secondary, not specified).

^f^ Patients with simultaneous diagnoses of venous thromboembolism and thrombocytopenia.

**Table S8.** **Venous thromboembolism in the Covid-19 cohort (2021, weeks 1 to 16): 21 day crude incidence of the selected diagnoses, by age and sex, n/100 000.**

| Age | n | VTE^a^ | CVST^b^ | MesT^c^ | PVT^d^ | TCP^e^ | VTE+TCP^f^ |
| --- | --- | --- | --- | --- | --- | --- | --- |
| 10–19 | 29 088 | 0 | 0 | 0 | 0 | 10.31 | 0 |
| 20–29 | 24 660 | 20.28 | 0 | 0 | 0 | 28.39 | 0 |
| 30–39 | 27 570 | 39.90 | 0 | 0 | 0 | 54.41 | 3.63 |
| 40–49 | 36 735 | 138.83 | 0 | 5.44 | 0 | 114.33 | 2.72 |
| 50–59 | 31 020 | 386.85 | 0 | 0 | 12.89 | 235.33 | 19.34 |
| 60–69 | 18 901 | 910.00 | 10.58 | 10.58 | 31.74 | 476.17 | 52.91 |
| 70–79 | 12 683 | 1 419.22 | 0 | 47.31 | 15.77 | 961.92 | 110.38 |
| 80–89 | 7 572 | 1 518.75 | 0 | 39.62 | 13.21 | 871.63 | 13.21 |
| >89 | 2 387 | 1 340.59 | 0 | 0 | 41.89 | 1 005.45 | 0 |
| **Total** | **190 616** | **359.89** | **1.05** | **6.82** | **7.34** | **231.88** | **17.31** |
| 10–39 | 81 318 | 19.68 | 0 | 0 | 0 | 30.74 | 1.23 |
| 40–69 | 86 656 | 395.82 | 2.31 | 4.62 | 11.54 | 236.57 | 19.62 |
| 70–89 | 20 255 | 1 456.43 | 0 | 44.43 | 14.81 | 928.17 | 74.06 |
| >89 | 2 387 | 1 340.59 | 0 | 0 | 41.89 | 1 005.45 | 0 |
| Sex | | | | | | | |
| Men | **92 072** | **481.15** | **1.09** | **8.69** | **10.86** | **317.14** | **28.24** |
| Women | **98 544** | **246.59** | **1.01** | **5.07** | **4.06** | **152.22** | **7.10** |

^a^ Any diagnosis of venous thromboembolism.

^b^ Cerebral venous sinus thrombosis.

^c^ Mesenteric thrombosis.

^d^ Portal vein thrombosis.

^e^ Thrombocytopenia (idiopathic, secondary, not specified).

^f^ Patients with simultaneous diagnoses of venous thromboembolism and thrombocytopenia.

**Table S9.** **Age and sex distribution of people vaccinated with each Covid-19 vaccine.**

| **First dose** | | | | | | |
| --- | --- | --- | --- | --- | --- | --- |
| Age group | Comirnaty® | | Spikevax® | | VaxZevria® | |
|  | Men  (n=369 792) | Women  (n=619 326) | Men  (n=33 875) | Women  (n=49 134) | Men  (n=265 169) | Women  (n=324 968) |
| 10−39 | 6.6% | 9.6% | 17.3% | 27.0% | 11.0% | 18.8% |
| 40−69 | 14.8% | 20.0% | 55.7% | 56.5% | 88.9% | 81.2% |
| 70−89 | 72.3% | 60.9% | 26.6% | 16.3% | 0.0% | 0.0% |
| >89 | 6.3% | 9.4% | 0.3% | 0.2% | 0.0% | 0.0% |
| All | 100% | 100% | 100% | 100% | 100% | 100% |
| **Second dose** | | | | | | |
|  | (n=196 159) | (n=389 772) | (n=14 442) | (n=22 227) |  |  |
| 10–39 | 11.7% | 14.6% | 20.6% | 33.7% |  |  |
| 40–69 | 26.0% | 30.4% | 57.1% | 55.9% |  |  |
| 70–89 | 51.9% | 41.7% | 21.8% | 10.0% |  |  |
| >89 | 10.4% | 13.3% | 0.4% | 0.3% |  |  |
| All | 100% | 100% | 100% | 100% |  |  |

**Table S10.** **Non adjusted excess event rate of any diagnosis of VTE in people vaccinated with the first and with the second dose of Covid-19 vaccines and in patients with Covid-19 (21 days following vaccination or following the diagnosis of Covid-19), by age and sex.**

| Age, sex | Vaccine first dose  (n/100 000 doses) | Vaccine second dose  (n/100 000 doses) | Covid−19  (n/100 000 patients) |
| --- | --- | --- | --- |
| 10–39 | 1.86 (−0.62−4.34) | 2.08 (−1.68−5.84) | 18.43 (8.79−28.08) |
| 40–69 | 6.63 (3.92−9.34) | 9.14 (3.40−14.88) | 388.66 (346.77−430.55) |
| 70–89 | 4.53 (0.21−8.85) | 10.46 (3.16−17.77) | 1 429.83 (1 263.63−1 596.03) |
| >89 | 6.58 (−7.44−20.60) | 2.33 (−11.79−16.45) | 1 306.98 (842.49−1 771.48) |
| Men | 19.00 (15.01−23.00) | 26.01 (18.05−33.96) | 472.50 (427.70−517.31) |
| Women | 9.02 (6.51−11.52) | 14.03 (9.58−18.47) | 239.50 (208.50−270.51) |
| **All** | **12.90 (10.71−15.10)** | **17.84 (13.86−21.83)** | **352.04 (325.11−378.97)** |

**Table S11.** **Non adjusted excess event rate (ER) estimates of unusual site thrombosis^a^ associated to thrombocytopenia per 100 000 first dose vaccinations, by age and sex.**

| Age group, sex | Excess rate (n/100 000) |
| --- | --- |
| 10−19 | −0.15 |
| 20−29 | −0.18 |
| 30−39 | 2.56 |
| 40−49 | 3.78 |
| 50−59 | 3.45 |
| 60−69 | 0.13 |
| 70−79 | 2.82 |
| 80−89 | −1.58 |
| >89 | −1.73 |
| **Total** | **3.50** |
| 10−39 | 1.33 |
| 41−69 | 2.49 |
| 70−89 | 0.81 |
| >89 | −1.73 |
| **Men** | 6.11 |
| **Women** | 1.89 |

^a^	Any of the following: cerebral venous sinus thrombosis, mesenteric thrombosis, portal vein thrombosis, or any venous thromboembolism associated with thrombocytopenia.

**Table S12.** **Venous thrombembolism (VTE) in unusual anatomical sites as a proportion of all cases of VTE in the four cohorts.**

|  | Any VTE, n | Unusual VTE or VTE+TCP, n | Percentage (95 percent confidence interval) |
| --- | --- | --- | --- |
| Reference | 9 539 | 2 619 | 27.5 (26.6−28.4) |
| Vaccinated first dose | 345 | 94 | 27.3 (22.8−32.2) |
| Vaccinated second dose | 160 | 45 | 28.1 (21.7−35.5) |
| Covid-19 | 686 | 62 | 9.0 (7.1−11.4) |

**Table S13**. **Number of cases and incidence of unusual site thrombosis^a^ and of simultaneous diagnosis of venous thromboembolism (VTE) and thrombocytopenia (TCP) in vaccinated individuals, by vaccine, following the first and following the second dose; 2021, weeks 1 to 16.**

|  |  | Unusual thrombosis or VTE+TCP | | Simultaneous diagnosis of VTE and TCP | |
| --- | --- | --- | --- | --- | --- |
|  | Doses, n | Cases, n | I, n/100 000 (95 percent confidence interval) | Cases, n | I, n/100 000 (95 percent confidence interval) |
| **First dose** | | | | | |
| Comirnaty® | 989 118 | 58 | 5.86 (4.53−7.58) | 6 | 0.61 (0.27−1.35) |
| Spikevax® | 83 009 | 19 | 22.89 (14.60−35.88) | 3 | 3.61 (1.17−11.21) |
| VaxZevria® | 590 137 | 17 | 2.88 (1.79−4.63) | 5 | 0.85 (0.35−2.04) |
| Total | **1 662 264** | **94** | 5.65 (4.62−6.92) | **14** | 0.84 (0.50−1.42) |
| **Second dose** | | | | | |
| Comirnaty® | 585 931 | 38 | 6.49 (4.72−8.91) | 4 | 0.68 (0.26−1.82) |
| Spikevax® | 36 669 | 7 | 19.09 (9.10−40.04) | 1 | 2.73 (0.38−19.36) |
| VaxZevria® | 178 | 0 | 0 | 0 | 0 |
| Total | **622 778** | **45** | 7.23 (5.39−9.68) | **5** | 0.80 (0.33−1.93) |

^a^	Any of the following: cerebral venous sinus thrombosis, mesenteric thrombosis, portal vein thrombosis, or any venous thromboembolism associated with thrombocytopenia.

**Table S14.** **Number of cases of various diagnostic definitions following the first and following the second dose of each vaccine; 2021, weeks 1 to 16.**

|  | Doses, n | Unusual thrombosis or VTE+ TCP^a^ | VTE+TCP^b^ | Any VTE^c^ | CVST^d^ | MesT^e^ | PVT^f^ | TCP^g^ |
| --- | --- | --- | --- | --- | --- | --- | --- | --- |
| **First dose** |  |  |  |  |  |  |  |  |
| Comirnaty® | 989 118 | 58 | 6 | 241 | 2 | 23 | 27 | 166 |
| Spikevax® | 83 009 | 19 | 3 | 68 | 0 | 4 | 12 | 77 |
| VaxZevria® | 590 137 | 17 | 5 | 36 | 4 | 3 | 5 | 22 |
| **All** | **1 662 719** | **94** | **14** | **345** | **6** | **30** | **44** | **265** |
| **Second dose** |  |  |  |  |  |  |  |  |
| Comirnaty® | 585 931 | 38 | 4 | 136 | 1 | 16 | 17 | 109 |
| Spikevax® | 36 669 | 7 | 1 | 24 | 0 | 2 | 4 | 24 |
| VaxZevria® | 178 | − | − | − | − | − | − | − |
| **All** | **622 778** | **45** | **5** | **160** | **1** | **18** | **21** | **133** |

^a^	Any of the following: cerebral venous sinus thrombosis, mesenteric thrombosis, portal vein thrombosis, or any venous thromboembolism associated with thrombocytopenia.

^b^	Patients with simultaneous diagnoses of venous thromboembolism and thrombocytopenia.

^c^	Any diagnosis of venous thromboembolism.

^d^	Cerebral venous sinus thrombosis.

^e^	Mesenteric thrombosis.

^f^	Portal vein thrombosis.

^g^ Thrombocytopenia (idiopathic, secondary, not specified).
